# Immunogenicity and safety of the homogenous booster shot of a recombinant fusion protein vaccine (V-01) against COVID-19 in healthy adult participants primed with a two-dose regimen

**DOI:** 10.1101/2021.11.04.21265780

**Authors:** Yuan Li, Xin Fang, Rongjuan Pei, Renfeng Fan, Shaomin Chen, Peiyu Zeng, Zhiqiang Ou, Jinglong Deng, Jian Zhou, Zehui Sun, Lishi Liu, Hua Peng, Xujia Chen, Zhipeng Su, Xi Chen, Jianfeng He, Wuxiang Guan, Zhongyu Hu, Yang-Xin Fu, Jikai Zhang

**Affiliations:** Guangdong Provincial Institute of Biological Products and Materia Medica, Guangzhou, China; Guangdong Provincial Center for Disease Control and Prevention, Guangzhou, China; National Institutes for Food and Drug Control, Beijing, China; Wuhan Institute of Virology, Chinese Academy of Sciences, Wuhan, China; Gaozhou Center for Disease Control and Prevention, Maoming, China; LivzonBio Inc., Zhuhai, China; Key Laboratory of Infection and Immunity, Institute of Biophysics, Chinese Academy of Sciences, Beijing, China; Guangzhou Laboratory, and Bioland Laboratory (Guangzhou Regenerative Medicine and Health Guangdong Laboratory), Guangzhou, China; Department of Basic Medical Sciences, School of Medicine, Tsinghua University, Beijing, China

**Keywords:** COVID-19, Recombinant fusion protein vaccine, Immunogenicity, Safety, SARS-CoV-2, VOCs

## Abstract

**Background:** Rising concerns over waning immunity and reduction in neutralizing activity against variants of concern (VOCs) have contributed to deploying booster doses by different strategies to tackle the COVID-19 pandemic. Preliminary findings from Phase I and II have shown that V-01, a recombinant fusion protein vaccine against COVID-19, exhibited favorable safety and immunogenicity profiles in 1060 adult participants of both younger and senior age. Herein, we aimed to assess the immunogenicity and safety for a booster dose in participants previously primed with a two-dose 10μg V-01 regimen (day 0, 21) from phase I trial, providing reassuring data for necessity and feasibility of a homogenous booster dose.

**Methods:** We conducted a single-arm, open-label trial at the Guangdong Provincial Center for Disease Control and Prevention (Gaozhou, China). Forty-three eligible participants who were previously primed 4-5 months earlier with two-dose 10μg V-01 regimen from phase I trial received booster vaccination. We primarily assessed the immunogenicity post-booster vaccination, measured by RBD-binding antibodies using ELISA and neutralizing activity against wild-type SARS-CoV-2 and emerging variants of concern (VOCs) using neutralization assays. We secondarily assessed the safety and reactogenicity of the booster vaccination.

**Results:** The third dose of V-01 exhibited significant boosting effects of humoral immune response in participants primed with two-dose 10μg V-01 regimen regarding both wild-type SARS-CoV-2 and VOCs. We observed a 60.4-folds increase in neutralizing titres against SARS-CoV-2 of younger adults, with GMTs of 17 (95%CI: 12-23) prior to booster vaccination in comparison to 1017 (95%CI: 732-1413) at day 14 post booster vaccination; and a 53.6-folds increase in that of older adults, with GMTs of 14 (95%CI: 9-20) before booster vaccination in comparison to 729(95%CI: 397-1339) at day 14 post-booster vaccination. The neutralizing titres against SARS-CoV-2 Delta strain also demonstrated a sharp increase from the day of pre booster vaccination to day 14 post booster vaccination, with GMTs of 11 (95%CI:8-15) versus 383 (95%CI:277-531) in younger adults (35.4-folds increase), and 6.5(95%CI: 5-8) versus 300(95%CI:142-631) in older adults (46.0-folds increase), respectively. We also observed a considerable and consistent increase of pseudovirus neutralizing titres against emerging VOCs from day 28 post second vaccination to day 14 post booster vaccination, with GMTs of 206 (95%CI:163-259) versus 607 (95%CI: 478-771) for Alpha strain, 54 (95%CI:38-77) versus 329 (95%CI: 255-425) for Beta strain, 219 (95%CI:157-306) versus 647 (95%CI: 484-865) for Delta strain. Our preliminary findings indicate a homogenous booster dose of V-01 was safe and well-tolerated, with overall adverse reactions being absent or mild-to-moderate in severity, and no grade 3 or worse AEs were related to booster vaccination.

**Conclusions:** A homogenous booster immunization in participants receiving a primary series of two-dose V-01 elicited a substantial humoral immune response against wild-type SARS-CoV-2 and emerging VOCs, along with a favorable safety and reactogenicity profile. Our study provided promising data for a homogenous prime-boost strategy using recombinant protein vaccine to tackle the ongoing pandemic, potentially providing broad protection against emerging VOCs and overcoming waning immunity.

## Introduction

Globally, there have been over 216 million confirmed cases of COVID-19, including 4.5 million deaths, according to the data reported to WHO, as of 30 August 2021. A total of more than 6 billion COVID-19 vaccine doses have been administered across over 190 countries. Seven vaccine candidates has been validated by WHO for emergency use, including BNT162b2 by Pfizer/BioNTech (mRNA), mRNA-1273 by Moderna (mRNA), Ad26.COV2.S by Janssen (adenovirus), AZD1222 by Oxford/AstraZeneca (adenovirus), Covishield by Serum Institute of India (adenovirus), BBIBP-CorV by Sinopharm Beijing (inactivated), CoronaVac by Sinovac (inactivated), and another 16 vaccines gained approval by at least one country, including ZF2001, Ad5-nCoV, Sputnik V, Sputnik Light, Covaxin, etc. The mRNA-based vaccines (mRNA-1273 and BNT162b2) can provide over 94% protective efficacy against symptomatic COVID-19 and elicit strong T-cell and B-cell immune responses, with mean neutralization antibody titer approximately 4 folds from that of convalescent panels. However broader usage in developing countries may be limited by the stringent storage and transportation conditions, relatively high cost, as well as concerns over the high incidence of adverse events (AEs). A single injection of adenovirus vector-based vaccines induces robust T cell responses but relatively lower humoral responses and should overcome pre-existing anti-vector immunity. Inactivated vaccines demonstrated promising safety profiles and good protective efficacy, while T-cell mediated immune responses are usually absent or very weak. The manufacturing capacity of such vaccines using wild-type SARS-CoV-2 strains was restrained by the biosafety level 3 and above facilities. Recombinant protein vaccines exhibited encouraging safety profiles and large scale manufacturing capacity, but need repeated doses to elicit robust immune responses.

Different strategies for booster shots have been scheduled in countries with high vaccination rates, considering vaccine properties, waning immunity, the prevalence of emerging VOCs. Substantial evidences have indicated the necessity of a booster immunization. Firstly, recent studies from Europe have assessed heterologous prime-boost immunization (a strategy also called *mix-and-match*) in Pfizer and AstraZeneca COVID-19 vaccines. The heterologous prime-boost vaccination regimen elicited significantly higher neutralization capacity than homologous ChAdOx1 or BNT162b2 immunization^1^. It is speculated that the mix-and-match strategies may elicit more potent and broader immune responses (including both humoral and cellular immune responses), which can also provide flexible choices to speed up vaccination campaigns worldwide for better control of the current pandemic. Additionally, as the dominant Delta variant (B.1.617.2), accounting for over 95% of the new SARS-CoV-2 infections, surges worldwide in the last few months, the number of SARS-CoV-2 new infections continues to rise, reversing what had been a decline since early 2021. Notably, real-world evidences have indicated reduced protective efficacy or neutralization activity for vaccines that gained official approval or emergency use authorization (EUA) against COVID-19^2-4^, which might be explained by the latest immune durability data exhibiting a slight expected decline in titres of binding and neutralizing antibodies^5^. These findings are consistent with the estimation of a statistical model that vaccine with an initial efficacy of 95% would expect to maintain efficacy of 77% after 250 days, while an initial efficacy of 70% may drop to a lower efficacy of 33% after 250 days, on the prerequisite that neutralization is the principal mechanism of protection^6^. Particular attention should be paid that even a considerable number of fully vaccinated individuals experienced breakthrough infections^7-9^. A booster exposure to antigen may awaken memory cells produced in the primary dose regimen, leading to a faster and stronger immune response (a process called *affinity maturation* for memory B cells). A study has shown that an additional dose of vaccine administered for 6 months or a longer interval after a second dose effectively recalled specific immune responses to SARS-CoV-2, resulting in a remarkable increase in antibody levels^10^. There have been substantial evidences that some SARS-CoV-2 variants acquired resistance to neutralizing antibodies generated by vaccination, reduction by a factor of several to tens of folds^11-13^. Nevertheless, it is assumed that high titer of neutralizing antibody is still correlated with protection, even in the presence of emerging VOCs that gained higher resistance to neutralization, and protection against severe COVID-19 cases remained preserved^14-17^. Ultimately, among immuno-compromised and immuno-suppressed populations, fold-reductions in antibody titres were observed after two mRNA vaccine doses^18-21^, which might result in persistent infection and accelerated mutation of SARS-CoV-2. The ability to substantially mount an immune response after an mRNA vaccine booster shot has been verified in solid organ transplant recipients^22^. Therefore, on 13 August, 2021, the Center for Disease Control and Prevention (CDC) of the United States recommended the people with moderately to severely immunocompromised conditions receive a booster shot of an mRNA COVID-19 vaccine after completion of the initial dose regimen.

Recently, rising concerns over waning immune response and reduction in neutralizing activity against variants of concern (VOC) have contributed to deploying booster shots^23^, including Russian, China, Brazil, the USA, and United Arab Emirates etc. CDC of the United States has announced that vaccine booster shots are available for vaccine recipients (completed their initial series for at least 6 months) aged over 65 years, or at-risk populations, such as living in long-term care settings, having underlying medical conditions, etc. The unequitable access (at-risk population remaining unvaccinated in low-income countries) to safe and effective vaccines across regions, absence of a reliable correlate of protection, inadequate data on performance of booster shots, and other considerations (e.g., optimal timing, boosting strategy and feasibility) might be the primary reasons for the moratorium on boosters by WHO until at least the end of 2021. Currently, evidences for prime-boost vaccination strategy remain sparse, although the reassuring data on the necessity of booster shots are trickling in. V-01, a recombinant fusion protein vaccine against COVID-19, in which RBD is armed with an interferon-α (IFN-α) at the N-terminus and dimerized by human IgG1 Fc at the C-terminus, with further addition of a pan HLA-DR-binding epitope (IFN-PADRE-RBD-Fc dimer), were designed as a strategic self-adjuvanted antigen with the conventional alum adjuvant^24^. Previous studies^25 26^ have shown that V-01 exhibited favorable safety profiles and provoked substantial immune responses in 1060 participants aged 18 years and older. An additional dose was inoculated in participants previously (4-5 months earlier) primed with the two-dose regimen of 10 μg V-01 (day 0, 21) in phase I trial. Herein, we present the preliminary immunogenicity and safety findings of the booster shot in fully vaccinated cohorts (two-dose 10 μg V-01) in phase I.

## Methods

### Study design and participants

We initiated a single-arm, open-label trial at the Guangdong Provincial Center for Disease Control and Prevention (Gaozhou, China). Eligible participants were those who completed the initial two-dose regimens of 10μg V-01 in phase I trial^25^ 4-5 months earlier and who did not meet the withdrawal criteria, voluntarily consented to participate in this trial, agreed to take effective contraceptive measures (women of childbearing potential) during the study, and without a history of SARS-CoV-2 infection or close contact to asymptomatic individuals. Exclusion criteria were: receipt of a COVID-19 vaccine other than V-01; hyperpyrexia (axillary temperature ≥ 39.0°C) lasting ≥3 days following a previous V-01 administration; severe or medically attended allergic reactions following a previous V-01 inoculation; newly diagnosed severe chronic diseases or pre-existing chronic diseases poorly controlled by medication following primary V-01 immunization (for participants ≥60 years of age); any confirmed or suspected immunosuppression or immunodeficiency state as determined by medical history (e.g. HIV, asplenia); recurrent severe infections and use of immunosuppressive drugs, except for topical steroids or short-term oral steroids within the past 6 months; pregnant or lactating females, or those who plan to become pregnant within 6 months after the booster dose; receipt of immunoglobulin and/or any blood products within 3 months prior to booster immunization; and any other conditions that, in the opinion of the investigator, might interfere with the assessment of vaccine safety and immunogenicity outcomes or pose additional risks. Written informed consent of each participant was obtained prior to the booster vaccination. The trial protocol was reviewed and approved by the Institutional Review Board of the Guangdong Provincial Center of Disease Control and Prevention. The trial was registered with ClinicalTrials.gov (NCT05050474) and was conducted in accordance with Good Clinical Practice and the *Declaration of Helsinki*.

### Randomization and masking

Blinding and masking were not applicable in this open-label, single-arm study. Each eligible participants will be assigned a unique number based on the sequential order of enrollment.

### Procedures

Each participant received an intramuscular (IM) booster injection of 10μg V-01 in participants who were primed 4-5 months earlier with a two-dose regimen of 10μg V-01 from phase I trial. Subjects were scheduled for follow-up visits till 6 months after booster vaccination.

On day 0, before the booster inoculation, participants were screened for eligibility by their demographic characteristics, physical examinations, medical history, vital signs (blood pressure, pulse, and temperature), urine pregnancy tests (female of childbearing potential only); and blood samples were collected for baseline immune response evaluation. After the booster injection, participants were monitored for 30min for potential prompt post-vaccination adverse reactions. In the subsequent 7 days following the booster shots, participants were instructed to complete a diary card to record any AEs experienced (solicited local/systemic AEs and unsolicited AEs), daily body temperatures as well as concomitant medications. Solicited local AEs included pain, pruritus, redness, swelling, rash, and induration; solicited systemic AEs included fever, diarrhea, constipation, dysphagia, anorexia, vomiting, nausea, myalgia, arthralgia, headache, cough, dyspnea, pruritus (non-injection site), skin and mucosa abnormity, acute allergic reaction, and fatigue. On day 7, participants returned to the study site for diary card submission and were instructed to document any unsolicited AEs occurring within 8-30d following the booster administration. AEs were categorized according to the Medical Dictionary for Regulatory Activities (MedDRA) terms, classified by System Organ Class (SOC) and Preferred Term (PT), and graded in accordance with *the Guidelines for Grading of Adverse Events in Clinical Trials of Prophylactic Vaccine* by the National Medical Products Administration (NMPA) of China.

Blood samples were collected to determine of RBD-binding antibody and neutralizing antibody titres against wild-type SARS-CoV-2 or emerging VOCs on days 0, 14, 28 post booster vaccination. Serum samples were stored at −20°C or below and underwent cold-chain transfer to the testing laboratory for immunogenic analysis. The RBD-binding IgG tests (by ELISA)^25 26^ and pseudovirus [a vesicular stomatitis virus (VSV) pseudovirus production system expressing the spike glycoprotein^27^] neutralizing assays (Supplementary Methods), including cross-neutralizing assays against emerging VOCs, were conducted by the National Institute for Food and Drug Control (Beijing, China). We also assessed neutralizing activities against live SARS-CoV-2 and VOCs using cytopathic effect (CPE) assay (Supplementary Methods), tested by the Wuhan Institute of Virology, Chinese Academy of Science.

### Outcomes

The study primarily evaluated the immunogenicity and secondarily assessed the safety and reactogenicity of the booster shots administered 4-5 months later in participants primed with two-dose 10μg V-01 regimen from the phase I trial. The immunogenicity outcomes were geometric mean titer (GMT), geometric mean increases (GMI) of the RBD-binding antibody, and SARS-CoV-2 neutralizing antibody. Additionally, cross-neutralization activity against emerging VOCs was assessed.

The safety outcomes were the counts and percentages of AEs (AEs experienced within at least 30 minutes, solicited local/systemic and unsolicited AEs within 0-7 days, and unsolicited AEs within 8-30 days following the booster shot).

### Statistical analysis

A total of 43 participants were recruited in this trial to provide a preliminary assessment of the immunogenicity, safety and reactogenicity of booster shots in primary vaccinated cohorts (two-dose 10μg V-01 regimen). We performed safety analysis in boost Safety Set (bSS), including all participants who received the booster dose. We conducted immunogenicity analysis in boost Per Protocol Set (bPPS), including participants who had completed the booster immunization and had completed pre- and post-immunization blood sampling with available antibody results. We present counts and percentages of AEs, including overall AEs, AEs related to vaccination, AEs classified as grade 3 or worse, AEs leading to participant’s withdrawal. Descriptive summary statistics (counts, percentages) will be provided for any AE. The GMTs with associated Clopper-Pearson 95% CIs for the neutralizing and RBD-binding antibodies against wild-type SARS-CoV-2 were calculated. The GMTs of cross-neutralizing antibodies along with 95% CIs were also calculated for each VOCs after booster immunization. We applied a χ^2^ test or Fisher’s exact test to analyze categorical data and a t-test to compare log-transformed antibody titres between groups. Analyses were conducted using SAS 9.4 (SAS Institute Inc., San Diego, CA, USA), then graphed using GraphPad Prism 8.0 (GraphPad Software Inc., San Diego, CA, USA). P value less than 0.05 was considered statistically significant.

## Results

In this trial (Fig. 1), 43 eligible participants primed with two-dose 10μg V-01 regimen were finally enrolled to receive a booster immunization 4-5 months after the second dose, including 23 participants in the younger adult cohort and 20 participants in the older adult cohort. One participant in the older adult cohort withdrew owing to the fact that continuation of the experiment, in the opinion of investigator, may result in injury to the participant. Participants were all Han Chinese regarding the ethnicity and presented with a mean age of 41.3 (ranging from 29 to 55) with 5/23 (21.7%) being male, and 66.8 (ranging from 61-76) with 13/20 (65.0%) being male in the younger and older adult cohort, respectively.

**Figure 1.**
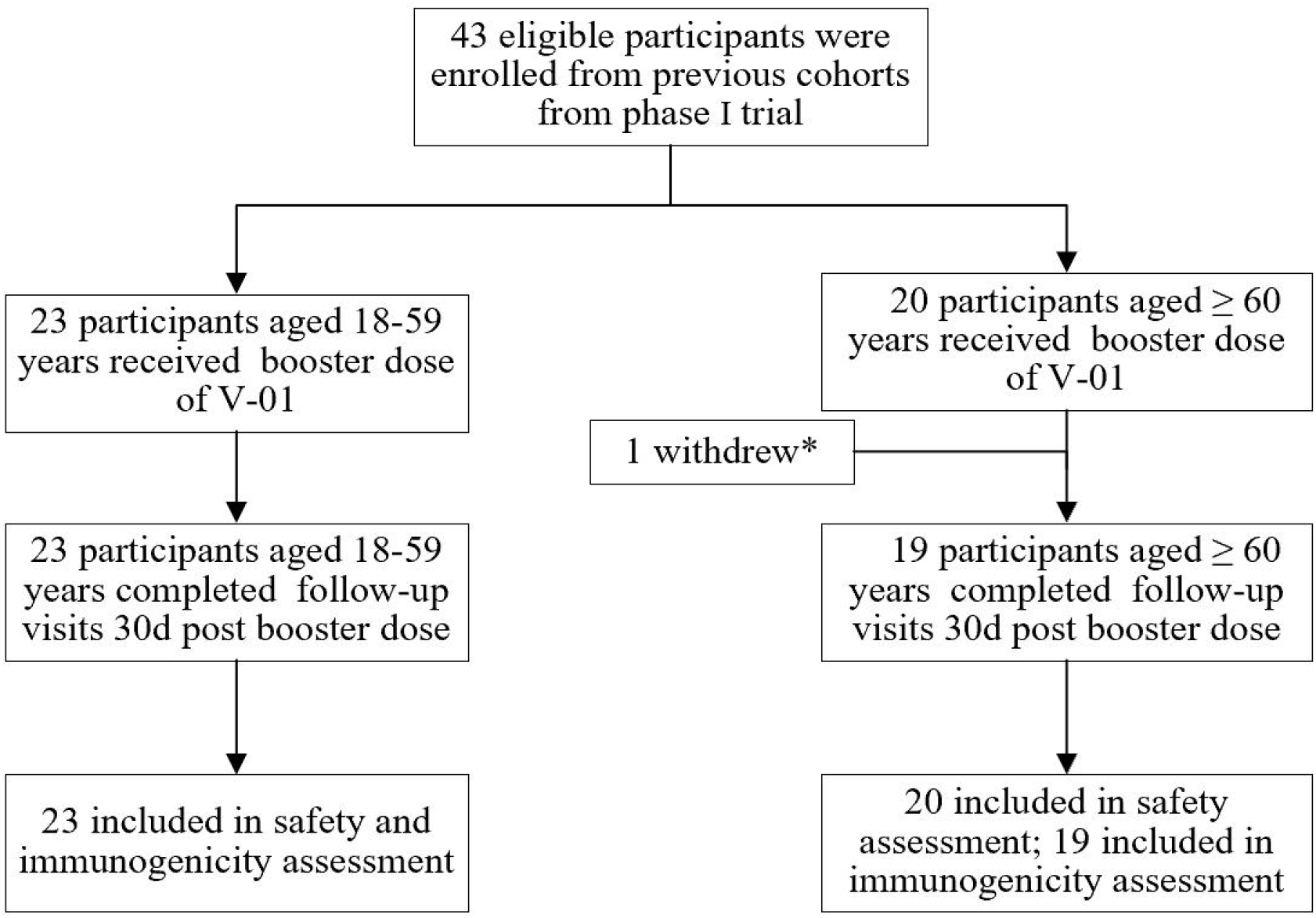
Trial flow of the booster vaccination. *One participant withdrew owning to experiencing grade 3 chronic gastritis which evaluated by the investigator that continuation of the experiment may result in injury to the participant.

A booster dose of V-01 exhibited significant boosting effects in terms of humoral immune response for participants primed with two-dose 10μg V-01 regimen regarding the RBD-binding IgGs against wild-type SARS-CoV-2. Following the booster dose, the RBD-binding antibody titres exhibited a dramatic increase at 14d, 28d post booster immunization compared with pre booster levels (Fig. 2A and 2B)), with GMTs of 4549(95%CI: 3374-6133), 3481(95%CI: 2645-4581) in comparison to 261 (95%CI: 194-352) in younger adults, 3562 (95%CI: 1805-7026), 3869(95%CI: 2235-6697) in comparison to 240(95%CI: 141-409) in older adults, and 251(95%CI: 189-333), 4060(95%CI: 2890-5702) in comparison to 3651(95%CI: 2769-4815) in merged cohorts, respectively.

**Figure 2.**
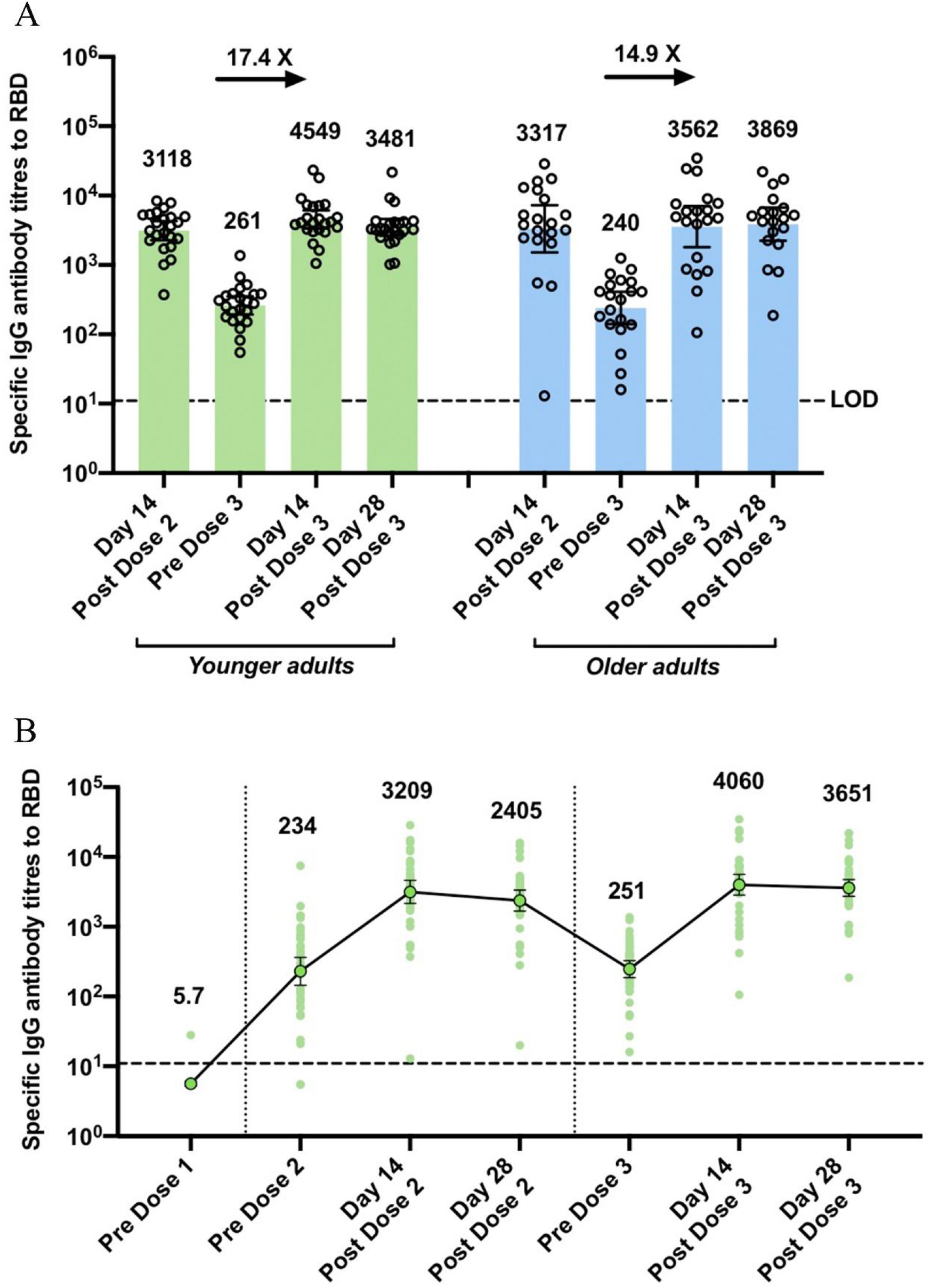
RBD-binding antibody titer against wild-type SARS-CoV-2 at pre- and post-booster vaccination. (A) Age stratified data; (B) Merged data since the initial dose.

Following the booster dose administration, the neutralizing antibody titres against wild-type SARS-CoV-2 exhibited a striking increase (Fig. 3A and 3B), which peaked at day 14 and remained at high levels at day 28 after the booster injection, with GMTs of 17 (95%CI: 12-23) at pre booster vaccination in comparison to 1017 (95%CI: 732-1413), 765 (95%CI: 608-962) at day 14, 28 post booster dose in younger adults, 14 (95% CI: 9-20) in comparison to 729 (95%CI: 397-1339), 697 (95%CI: 416-1165) in older adults, and 15 (95%CI: 12-19) in comparison to 871 (95%CI: 631-1202), 733 (95%CI: 570-943) in merged cohorts, respectively. The boosting effect was denoted by an amplification factor of 60.4 in younger adults, and 53.6 in older adults in comparison to the baseline neutralizing GMTs (before dose 3), and an amplification factor of 9.3 in younger adults, and 6.4 in older adults in comparison to the neutralizing GMTs of respective cohorts at day 14 after dose 2.

**Figure 3.**
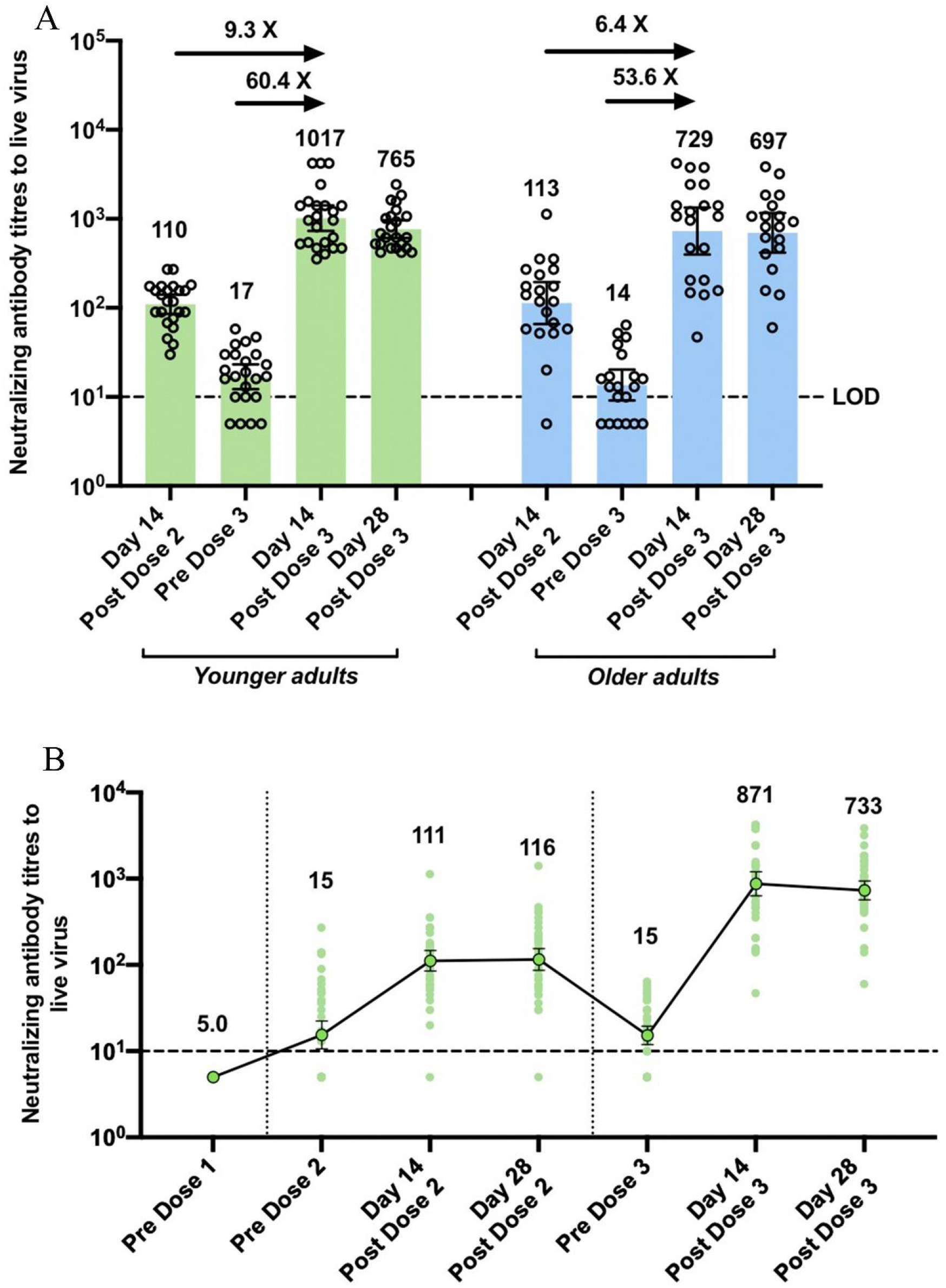
Neutralizing antibody titer against wild-type SARS-CoV-2 at pre- and post-booster vaccination. (A) Age stratified data; (B) Merged data since the initial dose.

We also observed a considerable and consistent increase pseudovirus neutralizing titres against emerging VOCs from day 28 post the second vaccination to day 14 post the booster vaccination (Fig. 4A and 4B), with GMTs of 206 (95%CI:163-259) versus 607 (95%CI: 478-771) for Alpha strain, 54 (95%CI:38-77) versus 329 (95%CI: 255-425) for Beta strain, 219 (95%CI:157-306) versus 647 (95%CI: 484-865) for Delta strain. Pseudovirus neutralizing antibody titres tended to reduce against SARS-CoV-2 VOCs (Alpha, Beta, Delta) in comparison to wild-type SARS-CoV-2 strain, whereas the titres remained fairly high enough to induce potential protection, particularly for those receiving booster shots.

**Figure 4.**
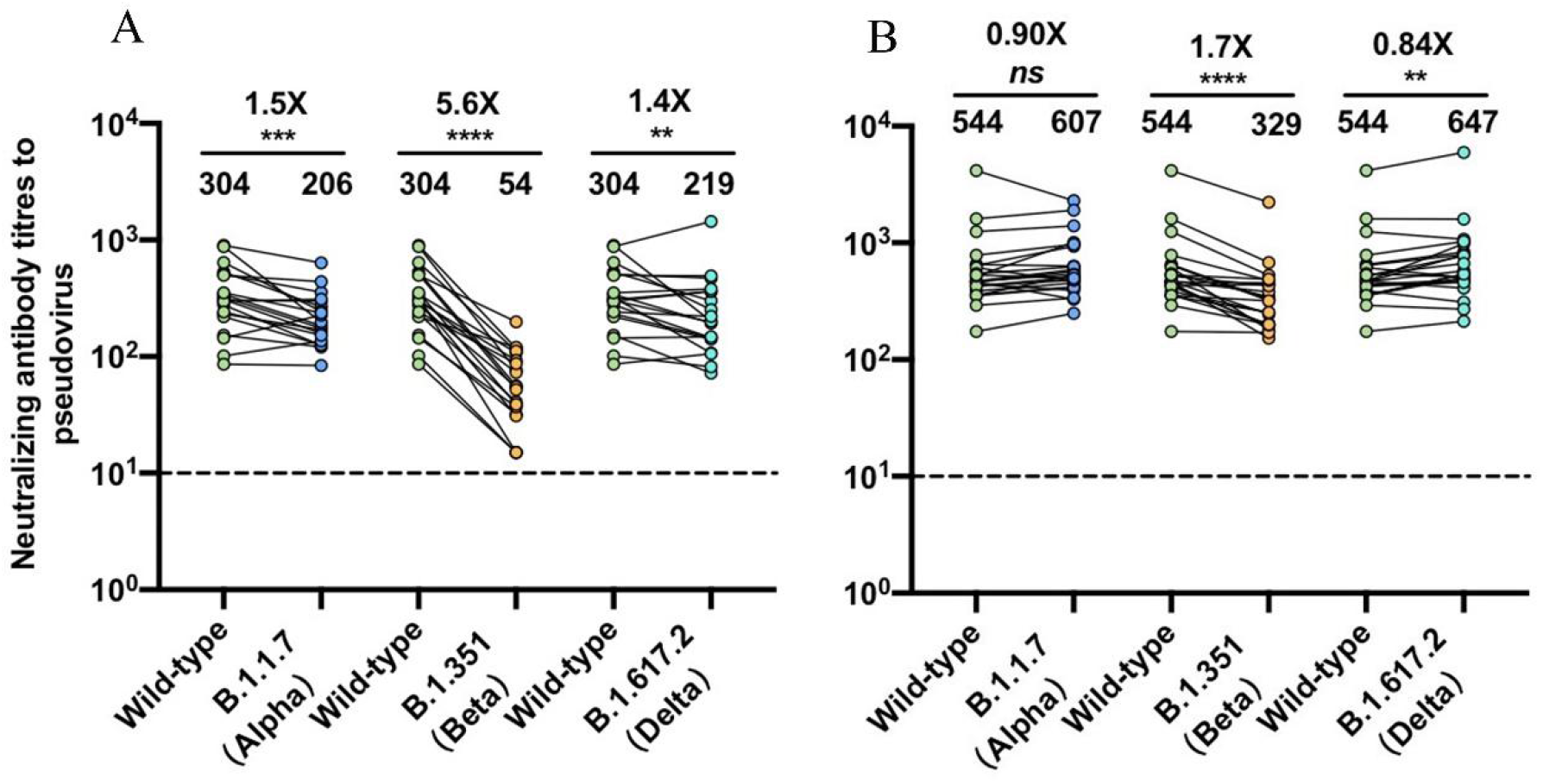
Cross-neutralization antibody titres against SARS-CoV-2 VOCs at (A) day 28 post the second immunization (from a sub-cohort of phase II), (B) day 14 post the booster immunization

Our preliminary safety results indicated that a booster dose of V-01 was safe and well-tolerated, with overall AEs being predominantly absent or mild-to-moderate in severity (Table 1). No grade 3 or worse AEs was attributable to the booster vaccination regarding both solicited and unsolicited AEs. We observed a quite acceptable overall local/systemic safety and reactogenicity profile within 7d post the homogenous booster immunization of V-01, with only one case of grade 2 arthralgia reported in the older adult cohorts and none reported in the younger adult group. As regard to unsolicited AEs, 2 cases (2/23, 8.7%) in the younger cohort, 1 case (1/20, 5.0%) in the older cohort were reported within 30d following the booster vaccination, but were all assessed as unrelated to the booster vaccination by the investigator.

**Table 1.** Overall adverse events, solicited local and systemic adverse reactions following booster dose of V-01 stratified by age

| Adverse events/reactions | Younger adults<br>(18-59 years, N=23) | Older adults<br>(≥60 years, N=20) |
| --- | --- | --- |
| **Overall adverse events within 30d following the booster dose** |  |  |
| Any | 0 | 2/20 (10.0) |
| Vaccination-related | 0 | 1/20 (5.0) |
| Grade 3 or worse | 0 | 0 |
| **Solicited local adverse reactions** |  |  |
| Pain | 0 | 0 |
| Induration | 0 | 0 |
| Swelling | 0 | 0 |
| Rash | 0 | 0 |
| Redness | 0 | 0 |
| Pruritus | 0 | 0 |
| **Solicited systemic adverse reactions** |  |  |
| Fever | 0 | 0 |
| Diarrhoea | 0 | 0 |
| Constipation | 0 | 0 |
| Dysphagia | 0 | 0 |
| Anorexia | 0 | 0 |
| Vomiting | 0 | 0 |
| Nausea | 0 | 0 |
| Myalgia | 0 | 0 |
| Arthralgia | 0 | 1/20 (5.0) |
| Headache | 0 | 0 |
| Cough | 0 | 0 |
| Dyspnoea | 0 | 0 |
| Pruritus | 0 | 0 |
| Skin and mucosa abnormality | 0 | 0 |
| Acute allergic reaction | 0 | 0 |
| Fatigue | 0 | 0 |
| **Unsolicited adverse events** |  |  |
| Any | 2/23 (8.7) | 1/20 (5.0) |
| Grade 3 or worse | 1/23 (4.4) | 1/20 (5.0) |
| Vaccination-related | 0 | 0 |

Data are presented as cases of adverse events/total participants in this cohort (%)

## Discussion

The safety and immunogenicity of V-01 have been previously assessed in a randomized, double-blind, placebo-controlled phase I trial using a dose-escalation and age-sequential design with a sentinel strategy, followed by the phase II trial with larger population of participants to further evaluate the immunogenicity and safety. Findings from phase I/II trial have substantiated a favorable immunogenicity and safety profile of V-01 both in younger adult and geriatric population, and advanced the two-dose, 10μg V-01 regimen to be tested in an international multicenter, large-scale phase III efficacy trial^25 26^. In this study, a homogenous booster dose in participants, who were primed 4-5 months earlier with two-dose 10μg V-01, not only elicited a potent boosting effect against both wild type and the dominant Delta SARS-CoV-2 strains but also showed a promising safety profile following V-01 booster.

Our preliminary findings provided reassurance that a homogenous booster shot of V-01 not only reversed the waning immunity to a steep increase in terms of antibody titres against SARS-CoV-2, but also elicited boosting effects against emerging VOCs. The humoral antibody levels faded gradually over time, judging by the fold-reduction of the antibody titer from day 28 post the second vaccination to the day of before booster vaccination, although RBD-binding antibody and neutralizing antibody in most of the subjects remained positive before booster vaccination. The degree of change for neutralizing antibody titres against live SARS-CoV-2 after the boosting dose as compared to second dose was much more significant than RBD-binding IgGs, indicating that V-01 booster dose could preferentially augment the functionality of neutralizing rather than increase quantity of total antibodies. Remarkably, at day 14 following the booster vaccination, neutralizing capacity against wild type and Delta strain of SARS-CoV-2 magnified by a factor of 60.4 versus 35.4 in younger adults, and 53.6 versus 46.0 in older adults, respectively. It is also noteworthy that neutralizing titer substantially increased from day 14 post dose 2 to day 14 post booster vaccination, with an augmentation factor of 9.3 in comparison to 2.4 in younger adults, and 6.4 in comparison to 2.1 in older adults, against wild-type and Delta strain of SARS-CoV-2 respectively. Meanwhile, the neutralizing capacity against emerging VOCs has been considerably promoted (2.9-6.1 folds) at 14d post booster dose in contrast to day 28 post second dose as suggested by the results of pseudovirus assay. Our results demonstrate that a booster dose of V-01 has high potential to provide broad protection against emerging VOCs, particularly the Delta strain. The booster shot might reactivated immune memory and promote a process of affinity maturation in B cells, in which antigen-specific B cells undergo somatic hypermutation and affinity-based selection, resulting in a sharp increase of antibodies titres with increased antibody affinity and diversity^28^. Our findings were consistent with a study that reported a booster(third) dose of ChAdOx1 induced significantly higher antibody titres, above the level that correlates with over 80% efficacy after the second dose^29^. A booster dose of BNT162b2 administered 7 to 9 months following the primary two-dose series demonstrated that a booster dose could prolong protection and increase the breadth of protection against VOCs^30^.

In our study, the safety and reactogenicity profile following a homogenous booster dose exhibited an encouraging local/systemic safety and reactogenicity profile within 7 days post the homogenous booster immunization of V-01, when compared to the safety results from phase I trial in same cohorts after receiving each dose of 10μg V-01^25^. Our findings are consistent with other studies using a homogenous prime-boost strategy. A booster dose of the inactivated vaccine *CoronaVac* resulted in a lower percentage of AEs than that observed in the previous phase I/II clinical trial^10 31^. A study also reported a well-tolerated safety profile that the third dose of ChAdOx1 demonstrated lower reactogenicity than after the initial dose^29^.

The study has several limitations: (1) the administration interval between the booster dose and the second dose all ranged within 4-5 months, which might need a broader spectrum of range and further stratification for roll-out timing for booster doses. From public health perspective, the optimal timing of booster shot after prime immunization should consider multiple factors, including the current prevalence of COVID-19, supply of COVID-19 vaccine, emerging VOCs epidemiology, etc. (2) cross-neutralizing panels at day 28 after second vaccination who received a two-dose 10μg V-01 regimen in phase II was different from the cohorts receiving a booster dose. As a consequence, the comparison of cross-neutralizing activities against variants between primary two-dose series and booster vaccination should be interpreted with caution. (3) the sample size is relatively small, which is insufficient to assess rare vaccine-related AEs. (4) The long-term safety data of current cohorts in phase I trial was terminated since the application of the booster shots, and the T-cell immunity has not yet been analyzed.

In conclusion, the booster dose of V-01 in participants previously primed with 21d-apart two-dose 10μg V-01 in phase I trial elicited potent humoral response against both wild-type and Delta strain of SARS-CoV-2 and exhibited a favorable and well-tolerated safety profile. Albeit a substantial decline of immunity was observed between 14 days post-second dosing and pre-booster dosing, humoral responses sharply increased by a magnification factor of approximately 60 times following a booster shot and a factor of about 40 times against the dominant Delta strain. Our findings substantiate the roll-out of homogenous booster strategy for vaccinated individuals after 4-5 months of primary 2-dose vaccination, at least for those receiving recombinant protein vaccine.

## Data Availability

All data produced in the present work are contained in the manuscript

## Acknowledgements

This study is partially supported by the Emergency Key Program of Guangzhou Laboratory, Grant No. EKPG21-21, and funded by LivzonBio Inc. China. We thank all participants who volunteered for this study.

## References

1. Tenbusch M, Schumacher S, Vogel E, et al. Heterologous prime-boost vaccination with ChAdOx1 nCoV-19 and BNT162b2. Lancet Infect Dis 2021;21(9):1212–13. doi: 10.1016/S1473-3099(21)00420-5 [published Online First: 2021/08/02]

2. Lopez Bernal J, Andrews N, Gower C, et al. Effectiveness of Covid-19 Vaccines against the B.1.617.2 (Delta) Variant. N Engl J Med 2021;385(7):585–94. doi: 10.1056/NEJMoa2108891 [published Online First: 2021/07/22]

3. Nanduri S, Pilishvili T, Derado G, et al. Effectiveness of Pfizer-BioNTech and Moderna Vaccines in Preventing SARS-CoV-2 Infection Among Nursing Home Residents Before and During Widespread Circulation of the SARS-CoV-2 B.1.617.2 (Delta) Variant - National Healthcare Safety Network, March 1-August 1, 2021. MMWR Morb Mortal Wkly Rep 2021;70(34):1163–66. doi: 10.15585/mmwr.mm7034e3 [published Online First: 2021/08/27]

4. Tenforde MW, Self WH, Naioti EA, et al. Sustained Effectiveness of Pfizer-BioNTech and Moderna Vaccines Against COVID-19 Associated Hospitalizations Among Adults - United States, March-July 2021. MMWR Morb Mortal Wkly Rep 2021;70(34):1156–62. doi: 10.15585/mmwr.mm7034e2 [published Online First: 2021/08/27]

5. Widge AT, Rouphael NG, Jackson LA, et al. Durability of Responses after SARS-CoV-2 mRNA-1273 Vaccination. N Engl J Med 2021;384(1):80–82. doi: 10.1056/NEJMc2032195 [published Online First: 2020/12/04]

6. Khoury DS, Cromer D, Reynaldi A, et al. Neutralizing antibody levels are highly predictive of immune protection from symptomatic SARS-CoV-2 infection. Nat Med 2021;27(7):1205–11. doi: 10.1038/s41591-021-01377-8 [published Online First: 2021/05/19]

7. Hacisuleyman E, Hale C, Saito Y, et al. Vaccine Breakthrough Infections with SARS-CoV-2 Variants. N Engl J Med 2021;384(23):2212–18. doi: 10.1056/NEJMoa2105000 [published Online First: 2021/04/22]

8. Schieffelin JS, Norton EB, Kolls JK. What should define a SARS-CoV-2 “breakthrough” infection? J Clin Invest 2021;131(12) doi: 10.1172/JCI151186 [published Online First: 2021/05/12]

9. Vignier N, Berot V, Bonnave N, et al. Breakthrough Infections of SARS-CoV-2 Gamma Variant in Fully Vaccinated Gold Miners, French Guiana, 2021. Emerg Infect Dis 2021;27(10) doi: 10.3201/eid2710.211427 [published Online First: 2021/07/22]

10. Pan H, Wu Q, Zeng G, et al. Immunogenicity and safety of a third dose, and immune persistence of CoronaVac vaccine in healthy adults aged 18-59 years: interim results from a double-blind, randomized, placebo-controlled phase 2 clinical trial. medRxiv 2021:2021.07.23.21261026. doi: 10.1101/2021.07.23.21261026

11. Planas D, Bruel T, Grzelak L, et al. Sensitivity of infectious SARS-CoV-2 B.1.1.7 and B.1.351 variants to neutralizing antibodies. Nat Med 2021;27(5):917–24. doi: 10.1038/s41591-021-01318-5 [published Online First: 2021/03/28]

12. Planas D, Veyer D, Baidaliuk A, et al. Reduced sensitivity of SARS-CoV-2 variant Delta to antibody neutralization. Nature 2021;596(7871):276–80. doi: 10.1038/s41586-021-03777-9 [published Online First: 2021/07/09]

13. Wu K, Werner AP, Koch M, et al. Serum Neutralizing Activity Elicited by mRNA-1273 Vaccine. N Engl J Med 2021;384(15):1468–70. doi: 10.1056/NEJMc2102179 [published Online First: 2021/03/18]

14. Liu C, Ginn HM, Dejnirattisai W, et al. Reduced neutralization of SARS-CoV-2 B.1.617 by vaccine and convalescent serum. Cell 2021;184(16):4220–36 e13. doi: 10.1016/j.cell.2021.06.020 [published Online First: 2021/07/10]

15. Sheikh A, McMenamin J, Taylor B, et al. SARS-CoV-2 Delta VOC in Scotland: demographics, risk of hospital admission, and vaccine effectiveness. Lancet 2021;397(10293):2461–62. doi: 10.1016/S0140-6736(21)01358-1 [published Online First: 2021/06/18]

16. Wall EC, Wu M, Harvey R, et al. Neutralising antibody activity against SARS-CoV-2 VOCs B.1.617.2 and B.1.351 by BNT162b2 vaccination. Lancet 2021;397(10292):2331–33. doi: 10.1016/S0140-6736(21)01290-3 [published Online First: 2021/06/07]

17. Liu J, Liu Y, Xia H, et al. BNT162b2-elicited neutralization of B.1.617 and other SARS-CoV-2 variants. Nature 2021;596(7871):273–75. doi: 10.1038/s41586-021-03693-y [published Online First: 2021/06/11]

18. Pimpinelli F, Marchesi F, Piaggio G, et al. Fifth-week immunogenicity and safety of anti-SARS-CoV-2 BNT162b2 vaccine in patients with multiple myeloma and myeloproliferative malignancies on active treatment: preliminary data from a single institution. J Hematol Oncol 2021;14(1):81. doi: 10.1186/s13045-021-01090-6 [published Online First: 2021/05/19]

19. Rabinowich L, Grupper A, Baruch R, et al. Low immunogenicity to SARS-CoV-2 vaccination among liver transplant recipients. J Hepatol 2021;75(2):435–38. doi: 10.1016/j.jhep.2021.04.020 [published Online First: 2021/04/24]

20. Roeker LE, Knorr DA, Thompson MC, et al. COVID-19 vaccine efficacy in patients with chronic lymphocytic leukemia. Leukemia 2021;35(9):2703–05. doi: 10.1038/s41375-021-01270-w [published Online First: 2021/05/15]

21. Shostak Y, Shafran N, Heching M, et al. Early humoral response among lung transplant recipients vaccinated with BNT162b2 vaccine. Lancet Respir Med 2021;9(6):e52–e53. doi: 10.1016/S2213-2600(21)00184-3 [published Online First: 2021/05/09]

22. Kamar N, Abravanel F, Marion O, et al. Three Doses of an mRNA Covid-19 Vaccine in Solid-Organ Transplant Recipients. N Engl J Med 2021;385(7):661–62. doi: 10.1056/NEJMc2108861 [published Online First: 2021/06/24]

23. Callaway E. COVID vaccine boosters: the most important questions. Nature 2021;596(7871):178–80. doi: 10.1038/d41586-021-02158-6 [published Online First: 2021/08/07]

24. Sun S, Cai Y, Song TZ, et al. Interferon-armed RBD dimer enhances the immunogenicity of RBD for sterilizing immunity against SARS-CoV-2. Cell Res 2021;31(9):1011–23. doi: 10.1038/s41422-021-00531-8 [published Online First: 2021/07/17]

25. Zhang J, Hu Z, He J, et al. Safety and immunogenicity of a recombinant interferon-armed RBD dimer vaccine (V-01) for COVID-19 in healthy adults: a randomized, double-blind, placebo-controlled, Phase I trial. Emerg Microbes Infect 2021;10(1):1589–97. doi: 10.1080/22221751.2021.1951126 [published Online First: 2021/07/02]

26. Shu YJ, He JF, Pei RJ, et al. Immunogenicity and safety of a recombinant fusion protein vaccine (V-01) against coronavirus disease 2019 in healthy adults: a randomized, double-blind, placebo-controlled, phase II trial. Chin Med J (Engl) 2021;134(16):1967–76. doi: 10.1097/CM9.0000000000001702 [published Online First: 2021/07/27]

27. Nie J, Li Q, Wu J, et al. Establishment and validation of a pseudovirus neutralization assay for SARS-CoV-2. Emerg Microbes Infect 2020;9(1):680–86. doi: 10.1080/22221751.2020.1743767 [published Online First: 2020/03/25]

28. Quast I, Tarlinton D. B cell memory: understanding COVID-19. Immunity 2021;54(2):205–10. doi: 10.1016/j.immuni.2021.01.014 [published Online First: 2021/01/30]

29. Flaxman A, Marchevsky NG, Jenkin D, et al. Reactogenicity and immunogenicity after a late second dose or a third dose of ChAdOx1 nCoV-19 in the UK: a substudy of two randomised controlled trials (COV001 and COV002). Lancet 2021 doi: 10.1016/S0140-6736(21)01699-8 [published Online First: 2021/09/05]

30. Falsey AR, Frenck RW, Jr., Walsh EE, et al. SARS-CoV-2 Neutralization with BNT162b2 Vaccine Dose 3. N Engl J Med 2021 doi: 10.1056/NEJMc2113468 [published Online First: 2021/09/16]

31. Zhang Y, Zeng G, Pan H, et al. Safety, tolerability, and immunogenicity of an inactivated SARS-CoV-2 vaccine in healthy adults aged 18-59 years: a randomised, double-blind, placebo-controlled, phase 1/2 clinical trial. Lancet Infect Dis 2021;21(2):181–92. doi: 10.1016/S1473-3099(20)30843-4 [published Online First: 2020/11/21]

