## Supplemental Table 1 for "Immunogenicity and safety of the homogenous booster shot of a recombinant fusion protein vaccine (V-01) against COVID-19 in healthy adult participants primed with a two-dose regimen"

Table S1. GMT and seroconversion rates of specific RBD-binding IgG and neutralizing antibodies at pre-and post- booster dose

| | RBD-IgG antibody | | Neutralizing antibodies to pseudovirus | | Neutralizing antibodies to live virus | |
|---|---|---|---|---|---|---|
| | **Younger adult group** | **Older adult group** | **Younger adult group** | **Older adult group** | **Younger adult group** | **Older adult group** |
| **Day 14 (Post Dose 2)** | | | | | | |
| **n** | 23 | 20 | 23 | 20 | 23 | 20 |
| **Seroconversion** | 23 (100%, 85-100) | 20 (100%, 83-100) | 23 (100%, 85-100) | 19 (95%, 75-100) | 23 (100%, 85-100) | 19 (95%, 75-100) |
| **GMT** | 3118 (2275-4274) | 3317 (1518-7249) | 196 (151-253) | 179 (111-289) | 110 (85-141) | 113 (66-195) |
| **Day 0 (Pre-Dose 3)** | | | | | | |
| **n** | 23 | 20 | 23 | 20 | 23 | 20 |
| **Seroconversion** | 23 (100%, 85-100) | 20 (100%, 83-100) | 20 (87%, 66-97) | 9 (45%, 23-68) | 19 (83%, 61-95) | 14 (70%, 46-88) |
| **GMT** | 261 (194-352) | 240 (141-409) | 49 (39-62) | 31 (24-40) | 17 (12-23) | 14 (9-20) |
| **Day 14** | | | | | | |
| **n** | 23 | 20 | 23 | 20 | 23 | 20 |
| **Seroconversion** | 23 (100%, 85-100) | 20 (100%, 83-100) | 23 (100%, 85-100) | 20 (100%, 83-100) | 23 (100%, 85-100) | 20 (100%, 83-100) |
| **GMT** | 4549 (3374-6133) | 3562 (1805-7026) | 544 (414-715) | 509 (311-833) | 1017 (732-1413) | 729 (397-1339) |
| ***P* value** | 0.4588 | | 0.7677 | | 0.9445 | |
| **Day 28** | | | | | | |
| **n** | 23 | 19 | 23 | 19 | 23 | 19 |
| **Seroconversion** | 23 (100%, 85-100) | 19 (100%, 82-100) | 23 (100%, 85-100) | 18 (95%, 74-100) | 23 (100%, 85-100) | 19 (100%, 82-100) |
| **GMT** | 3481 (2645-4581) | 3869 (2235-6697) | 521 (418-650) | 492 (296-818) | 765 (608-962) | 697 (416-1165) |

Data are GMT (95% CI), number of participants (%, 95% CI) for seroconversion and *P* value of unpaired *t* test (younger adults GMT v.s. older adults GMT at Day 14).

Supplementary Methods

### SARS-CoV-2 pseudovirus neutralization assay

Pseudovirus neutralization assay was performed using the VSV-based luciferase reporter SARS-CoV-2 pseudoviruses bearing wild-type, B.1.1.7, B.1.351, and B.1.617.2 variants spike protein, which were provided by Beijing Tiantan Pharmaceutical Biotechnology Development Co. Ltd. The SARS-CoV-2 pseudotyped virus neutralization test began with 3-fold serially diluted heat-inactivated serum samples, which started at 1:5 and then mixed with a certain amount 650 $TCID_{50}$ of pseudotyped virus for about 1h. Then the mixture was added to Huh-7 cells ($2.5\times10^4$ per well). After that, the target cells were incubated for 24 h, and the amount of pseudotyped virus entering the target cells was calculated by detecting the expression of luciferase, to obtain the neutralizing antibody content of the sample. Following this protocol, four samples could be detected simultaneously in a 96-well plate. The cell control (CC) with only cells and the virus control (VC) with virus and cells were set up in each plate. When the raw data for control and samples were exported from the luminometer and pasted into the calculation template, and the neutralizing titer was calculated by the dilution number of 50% protective condition using Reed and Muench method. Samples with values ≥ 30 were defined as seropositive.

### Live SARS-CoV-2 virus amplification and titration

SARS-CoV-2 virus (wild-type strain collection No.: IVCAS 6.7512, Delta variant collection No.: IVCAS 6.7585) was propagated on Vero E6 cells. The virus was grown until the cytopathic efficiency (CPE) reached >75% and then harvested. The virus titer was determined by using a CPE assay as follows: $1\times10^4$ cells/well were seeded in a 96-well culture plate for 18–24 h, after which 10-fold serially diluted virus was added. Six repeats were included for each of six dilutions. Cells were cultured in a 5% $CO_2$ incubator at 37 °C and checked under a microscope for the presence of CPE after 4–5 days. The virus titer was calculated with the Reed and Muench method.

### Live SARS-CoV-2 neutralization assay

A CPE assay was used to determine the 50% neutralization titer to live SARS-CoV-2. Each serum sample was first incubated at 56 °C for 30 min for safety. Vero E6 cells were seeded in a 96-well culture plate for 18–24 h at a density of $1\times10^4$ cells/well. On the next day, the inactivated serum was serially diluted 3-fold, starting at 1:5, and six repeats were used for each dilution. In each well of the 96-well plate, 70 µl of serially diluted sera were mixed with 70 µl of 140 $TCID_{50}$ virus, then the sera/virus mixture was incubated at 37 °C (5% $CO_2$) for 2 h before transferring 100 µl of the sera/virus mixture to 96-well titer plates with confluent Vero E6 cells. After a 4-day incubation, the plate was observed under a microscope and the CPE of each well was recorded. The neutralizing titer was set as the dilution number of the 50% protective condition using the Reed and Muench method.
